# Metabolomics insights into osteoporosis through association with bone mineral density

**DOI:** 10.1101/2021.01.16.21249919

**Authors:** Xiaoyu Zhang, Hanfei Xu, Gloria H. Y. Li, Michelle T. Long, Ching-Lung Cheung, Ramachandran S. Vasan, Yi-Hsiang Hsu, Douglas P. Kiel, Ching-Ti Liu

**Affiliations:** Department of Biostatistics, Boston University School of Public Health, Boston, MA, 02118, USA; Department of Health Technology and Informatics, the Hong Kong Polytechnic University, Hung Hom, Hong Kong, China; Section of Gastroenterology, Boston Medical Center, Boston University School of Medicine, Boston, MA, 02118, USA; National Heart, Lung, and Blood Institute’s and Boston University’s Framingham Heart Study, Framingham, MA 01702, USA; Department of Pharmacology and Pharmacy, the University of Hong Kong, Pokfulam, Hong Kong, China; National Heart, Lung, and Blood Institute and Boston University’s Framingham Heart Study, Framingham MA 01702, USA; Section of Preventive Medicine and Epidemiology, Evans Department of Medicine, Boston University School of Medicine, Boston, MA 02118, USA; Whitaker Cardiovascular Institute and Cardiology Section, Evans Department of Medicine, Boston University School of Medicine, Boston, MA 02118, USA; Hinda and Arthur Marcus Institute for Aging Research, Hebrew SeniorLife, Boston, MA 02131, USA; Department of Medicine, Beth Israel Deaconess Medical Center and Harvard Medical School, Boston, MA 02215, USA; Broad Institute of MIT and Harvard, Boston, MA 02142, USA

**Author notes:** Correspondence to (CTL) and (XZ).

**Keywords:** General population studies, Osteoporosis, DXA, Fracture risk assessment Metabolomics

## Abstract

Osteoporosis, a disease characterized by low bone mineral density (BMD), increases the risk for fractures. Conventional risk factors alone do not completely explain measured BMD or osteoporotic fracture risk. Metabolomics may provide additional information. We aim to identify BMD-associated metabolomic markers that are predictive of fracture risk. We assessed 209 plasma metabolites by LC-MS/MS in 1,552 Framingham Offspring Study participants, and measured femoral neck (FN) and lumbar spine (LS) BMD 2-10 years later using dual energy x-ray absorptiometry. We assessed osteoporotic fractures up to 27-year follow-up after metabolomic profiling. We identified twenty-seven metabolites associated with FN-BMD or LS-BMD by LASSO regression with internal validation. Incorporating selected metabolites significantly improved the prediction and the classification of osteoporotic fracture risk beyond conventional risk factors (AUC=0.74 for the model with identified metabolites and risk factors vs AUC=0.70 with risk factors alone, p=0.001; Net reclassification index=0.07, p=0.03). We replicated significant improvement in fracture prediction by incorporating selected metabolites in 634 participants from the Hong Kong Osteoporosis Study (HKOS). The glycine, serine, and threonine metabolism pathway (including four identified metabolites: creatine, dimethylglycine, glycine, and serine) was significantly enriched (FDR p-value=0.028). Furthermore, three causally related metabolites (glycine, Phosphatidylcholine [PC], and Triacylglycerol [TAG]) were negatively associated with FN-BMD while PC and TAG were negatively associated with LS-BMD through Mendelian randomization analysis. In summary, metabolites associated with BMD are helpful in osteoporotic fracture risk prediction. Potential causal mechanisms explaining the three metabolites on BMD are worthy of further experimental validation. Our findings may provide novel insights into the pathogenesis of osteoporosis.

## Introduction

Osteoporosis is the most common metabolic bone disease, mainly characterized by low bone mineral density (BMD) and deteriorated bone strength and is associated with an increased risk of low-trauma fractures.^(1)^ Osteoporotic fractures, particularly vertebral and hip fractures, can be associated with chronic disabling pain and have a profound impact on patients’ quality of life.^(2)^ More than 75 million people in the United States, Europe, and Japan are affected by osteoporosis, which causes more than 8.9 million fractures annually worldwide.^(3)^ Therefore, prevention and early detection of osteoporosis are essential for people to maintain bone health and improve their overall quality of life.

Other than BMD, which is used for the diagnosis of osteoporosis, many clinical risk factors have been identified for osteoporotic fracture prediction such as age, female sex, premature menopause, smoking.^(4)^ A fracture risk assessment tool (FRAX), for example, has been developed based on the clinical risk factors to predict fracture.^(5)^ However, previous studies have shown that the aforementioned clinical risk factors do not completely explain measured BMD or fracture risk.^(6,7)^ Metabolomics, as products of metabolism, may provide additional information to predict BMD or may identify those individuals with a high probability of experiencing an osteoporotic facture.

Metabolites are small molecules that can be reactants, intermediates, or products of metabolism. Many previous studies show that metabolites are closely related to bone health.^(8-13)^ For example, a cross-sectional study identified a group of metabolites for characterizing low BMD in postmenopausal women.^(8)^ Another study also reported that metabolites represented useful markers to predict bone loss in menopausal women.^(9)^ However, these studies were limited by their relatively small sample sizes or a focus mainly on women. A larger community-based study including both women and men is needed to investigate potential relationships between metabolites and osteoporosis in the general population. Metabolomics may also provide insights into the biological mechanism of osteoporosis. In this study, our primary aim was to identify BMD-associated metabolomic markers that are predictive of fracture risk and to explore potential causal mechanisms between identified metabolites and BMD as a secondary goal.

## Subjects and Methods

### Discovery cohort

Framingham Heart Study (FHS) Offspring participants, aged 30-82 years, whose underwent plasma metabolite profiling at their fifth examination cycle 1991-1995 (baseline) and who underwent both femoral neck (FN) and lumbar spine (LS) BMD measurements between their sixth and the seventh examination cycles (1996-2001) comprised the sample for the present investigation. The FHS is a community-based cohort study that started in 1948, with the recruitment of 5,209 men and women between the ages of 28 and 62 years from Framingham, MA, USA.^(14)^ The FHS Offspring cohort enrolled the children of the original cohort and the children’s spouses, including 5,124 participants who underwent physical examination, medical history, and routine laboratory tests approximately every four years since 1971.^(15)^ There were 2067 Offspring participants with metabolites measurements at the fifth examination and 1604 of them also had BMD measurements between the sixth and the seventh examination. Individuals without body mass index information, smoking status, or women without menopausal status at the fifth examination were excluded. A total of 1552 individuals were eligible for the present investigation. All participants provided written informed consent and the study protocol was approved by the Hebrew SeniorLife and Boston University Medical Campus institutional review boards.

### Metabolite profiling

Plasma samples of FHS Offspring participants were collected in a fasting state at their fifth examination cycle between 1991 and 1995. High throughput metabolite profiling was performed on the collected plasma samples and the concentration of plasma metabolites was assessed using a liquid chromatography / mass spectrometry (LS/MS) platform as previously described.^(16,17)^ We removed metabolites with a high missing rate (≥ 20%) and replaced the missing values in the remaining metabolites by the half of minimum value of the same metabolite.^(18)^ A total of 209 metabolites were included in this study.

### BMD measurement and ascertainment of fracture incident

We measured BMD (g/cm^2^) at the femoral neck (greater trochanter & Ward’s area) and lumbar spine (average BMD of L2-L4) using a Lunar dual-energy x-ray absorptiometry (DXA) between the sixth and the seventh examination of the FHS Offspring cohort. Further details about BMD measurements were well described previously.^(19)^ Major incident osteoporotic fractures were assessed using self-report and subsequent medical record confirmation over 27 years of follow-up after the fifth examination. We defined incident osteoporotic fractures as follows: at least one fracture associated with falling from a standing height or less, excluding fracture at toes, fingers, skull, or face.

### Clinical covariates

Clinical covariates including age (years), sex, body mass index (BMI, kg/m^2^), current smoking status, and menopausal status (women only) were assessed at baseline (i.e. the fifth examination). BMI was calculated as weight divided by height squared. Current smoking status was defined as “yes” if an individual reported current smoking cigarettes regularly over the year preceding the Heart Study visit and “no” if an individual reported never smoking or smoking previously. Menopausal status in women was classified as “yes” for women whose menstruation stopped for at least 12 months and “no” otherwise. We additionally considered diabetes status, drinking status, and calcium intake at baseline for sensitivity analysis. Diabetes status was “yes” if an individual was treated for diabetes and “no” otherwise. Drinking status was defined as “yes” if an individual reported weekly drinking any type of alcohol and “no” otherwise. Calcium intake was “yes” if an individual reported taking calcium supplement and “no” otherwise.

### Replication cohort

The Hong Kong Osteoporosis Study (HKOS) is a prospective cohort established since 1995. The cohort profile has been described elsewhere.^(20)^ In brief, approximately 9,449 community-dwelling Southern Chinese subjects were recruited at baseline. Since 2015, these participants were invited to attend in-person follow-up visits. At both baseline and follow-up visits, the subjects were required to complete clinical assessments comprising physical examination (including measurement of height, weight, DXA-measured BMD, etc.) and self-reported questionnaires (such as tobacco use, menopause status, etc.). Fasting blood samples were also collected, with multiple aliquots of serum and plasma stored at −80 ^0^C. Part of the serum samples collected at baseline and in-person follow-up visits were sent to a liquid chromatography–mass spectrometry (LC-MS) platform, Metabolon, for untargeted metabolomic profiling.^(21)^ A total of 1,194 serum metabolites were profiled. Moreover, “in-silico” follow-up of the HKOS participants were performed using the electronic medical records available in the Clinical Data Analysis and Reporting System managed by the Hong Kong Hospital Authority.

### Statistical analyses

The clinical characteristics of participants are described by mean (SD) or count (%) in **Table 1**. Measurements of 209 plasma metabolites were natural logarithmically transformed and standardized. The flow-chart of analysis of this study is shown in **Fig. 1**. We first randomly split the entire sample into a testing dataset and a training dataset at a ratio of 4:6 by individuals’ family identification code to ensure that participants in the training dataset were unrelated to those in the testing dataset. For the training dataset, we then implemented a least absolute shrinkage and selection operator (LASSO) to select BMD-associated metabolites with tenfold cross validation, while the regularized parameter of LASSO with minimum mean cross-validated error was chosen.^(22)^ We compared three models, including a model with conventional risk factors, consisting of age, sex, BMI, current smoking status, and menopausal status, a model with selected metabolites alone, and a model with both conventional risk factors and selected metabolites, on the testing dataset using adjusted R-squared. We repeated the process described above 100 times and selected the metabolites with cumulative selection frequency greater or equal to 50. We conducted the analysis for both FN-BMD and LS-BMD individually and estimated the variation of BMD explained by the selected metabolites. We also performed a sex-stratified analysis as a secondary analysis. We calculated the average of the adjusted R-squared using the testing dataset for 100 iterations and the overall adjusted R-squared using the entire dataset based on the final selected metabolites to determine whether incorporation of the selected metabolites improved model fitting for BMD. We used the F-test to test whether two nested models (model with conventional risk factors alone vs model with conventional risk factors + selected metabolites) were significantly different. Finally, we compiled a list of overall selected metabolites which were selected by either FN-BMD or LS-BMD analyses.

**Table 1.**
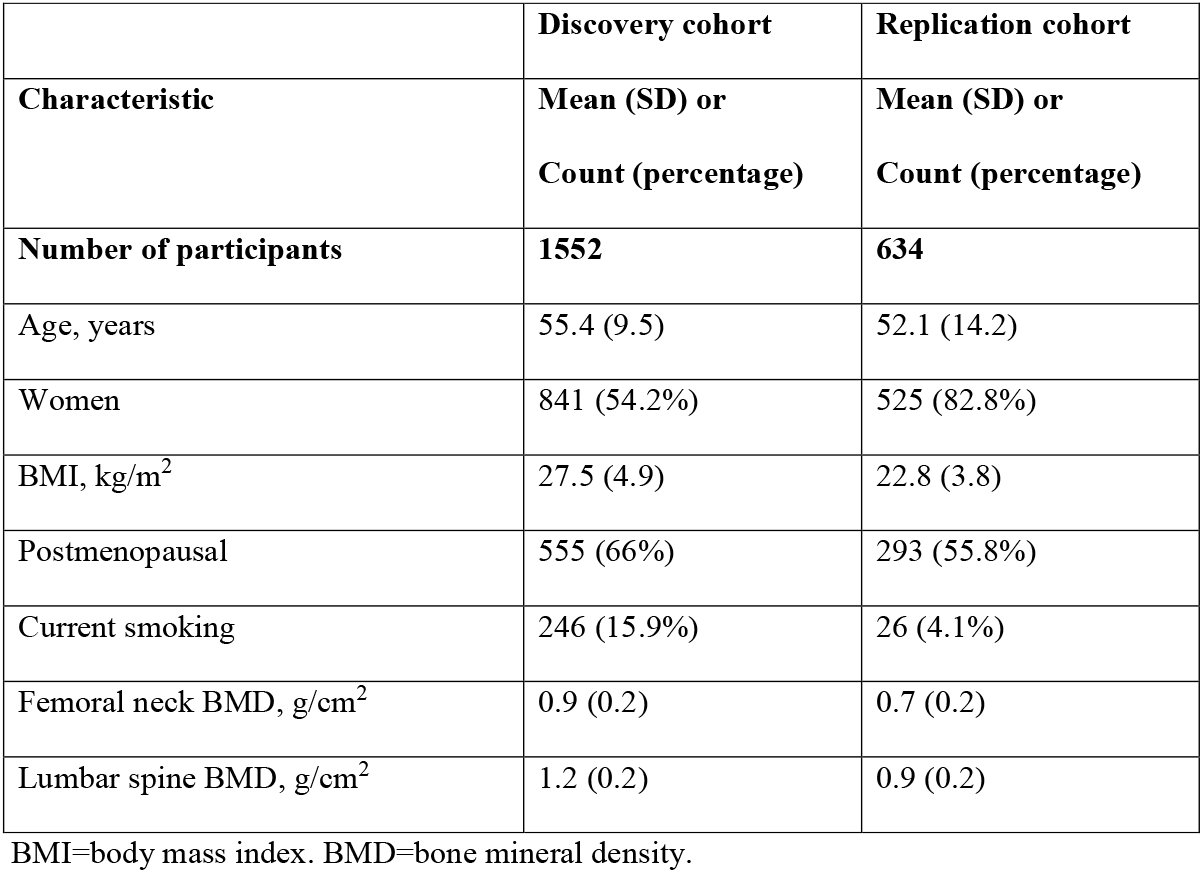
Clinical characteristics of participants in discovery and replication cohorts. Continuous variables are summarized as mean (SD=standard deviation) and categorical variables are summarized as count (percentage).

|  | **Discovery cohort** | **Replication cohort** |
| --- | --- | --- |
| **Characteristic** | **Mean (SD) or**<br>**Count (percentage)** | **Mean (SD) or**<br>**Count (percentage)** |
| **Number of participants** | **1552** | **634** |
| Age, years | 55.4 (9.5) | 52.1 (14.2) |
| Women | 841 (54.2%) | 525 (82.8%) |
| BMI, kg/m<sup>2</sup> | 27.5 (4.9) | 22.8 (3.8) |
| Postmenopausal | 555 (66%) | 293 (55.8%) |
| Current smoking | 246 (15.9%) | 26 (4.1%) |
| Femoral neck BMD, g/cm<sup>2</sup> | 0.9 (0.2) | 0.7 (0.2) |
| Lumbar spine BMD, g/cm<sup>2</sup> | 1.2 (0.2) | 0.9 (0.2) |

BMI=body mass index. BMD=bone mineral density.

**Fig. 1.**
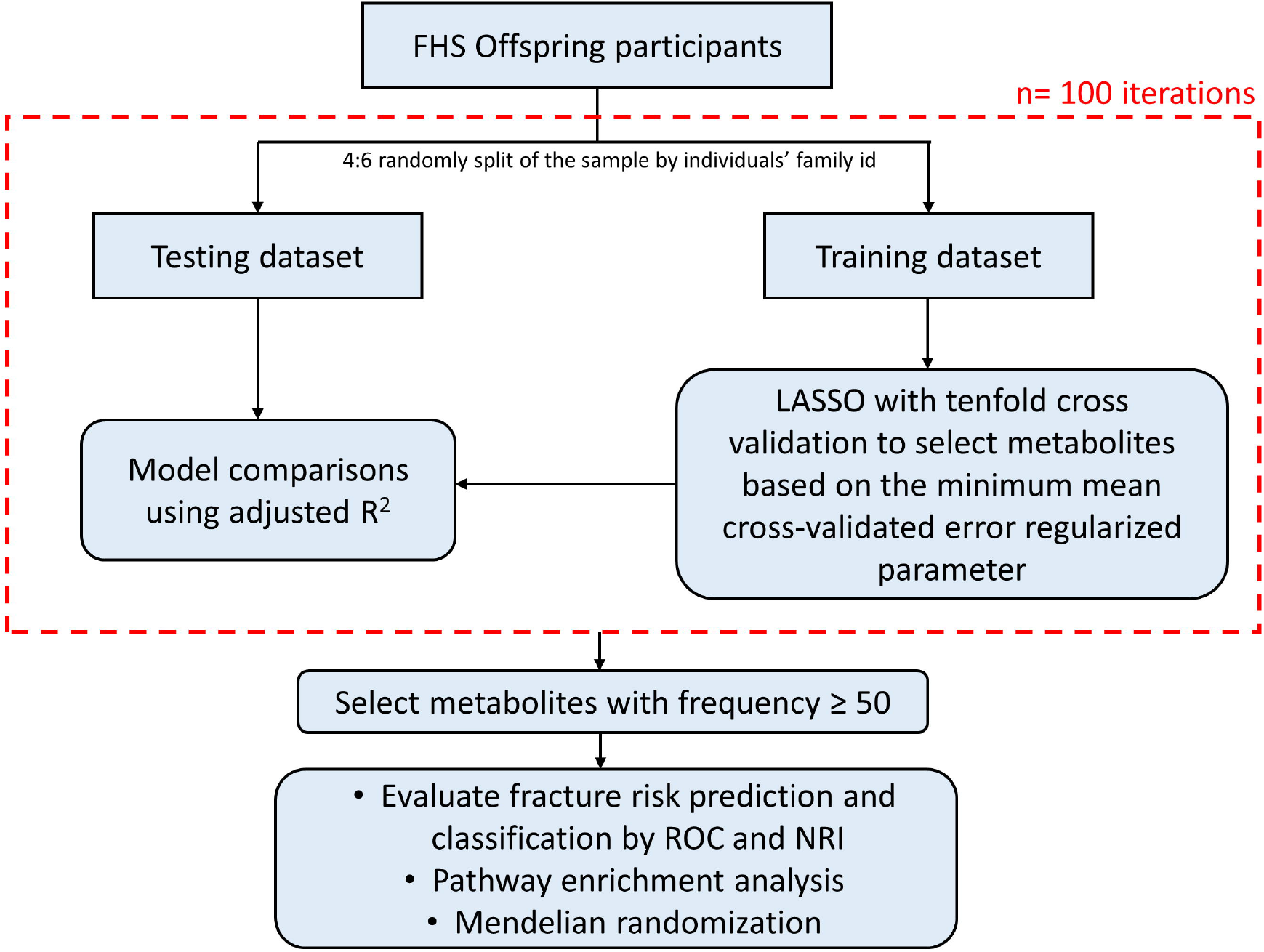
Flow-chart illustrating the main steps of the analysis. First, the entire sample was randomly split into a testing dataset and a training dataset at a ratio of 4:6 by individuals’ family id. For the training dataset, we then implemented a least absolute shrinkage and selection operator (LASSO) to select bone mineral density (BMD)-associated metabolites with ten-fold cross validation, while the regularized parameter of LASSO with minimum mean cross-validated error was chosen. Models with conventional risk factors alone, selected metabolites alone, and both conventional risk factors and selected metabolites were compared on the testing dataset through adjusted R-squared. We iterated the process (red dashed line) for 100 times and selected the metabolites with cumulative selection frequency greater or equal to 50. We conducted the analysis using both femoral neck (FN)-BMD and (LS)-BMD individually. Finally, we got a list of overall selected metabolites which were selected by either FN-BMD or LS-BMD. Next, we evaluated the fracture prediction accuracy and fracture risk classification using the overall selected metabolites on the entire dataset by the receiver-operating-characteristic (ROC) curve and net reclassification index (NRI). Pathway enrichment analysis was performed on the selected metabolites. We also conducted a two-sample Mendelian randomization for causal inference between selected metabolites and BMD.

In order to investigate whether the overall selected metabolites improved the prediction accuracy of osteoporotic fracture, we fitted a logistic regression model for osteoporotic fracture (outcome) on conventional risk factors and conventional risk factors plus overall selected metabolites (predictor variables) separately and then compared their performance for predicting the risk of fractures using the areas under the receiver-operating-characteristic (ROC) curve (AUC) on the entire dataset. Nonparametric test (DeLong’s test) was used to compare the discriminative capability of two correlated ROC curves.^(23)^ Net reclassification index (NRI) and integrated discrimination improvement (IDI) were also estimated to evaluate the improvement in model performance introduced by the inclusion of selected metabolites.^(24)^ Reclassification tables for subjects who do and do not experience major osteoporotic fracture during the follow-up were constructed into two categories using [0, 20%), [20%, 1] of predicted probability of fracture as the proxy of fracture risk based on the guideline of the National Osteoporosis Foundation of the United States.^(25)^ All p-values are two-sided and the level of significance was set to 0.05. We also added either FN-BMD or LS-BMD to the above two models and compared their fracture risk prediction and classification using ROC curves, NRI, and IDI. As a sensitivity analysis, we included diabetes status, drinking status, and calcium intake as additional covariates in the model and repeated the same statistical strategies described above.

Pathway enrichment analysis was performed on the selected metabolites using MetaboAnalyst 4.0 (https://www.metaboanalyst.ca/).^(26)^ The false discovery rate (FDR) of 0.05 was set as the significance level for pathway enrichment analysis. Next, we conducted a two-sample Mendelian randomization for causal inference for selected metabolites associated with BMD. We considered genetic variants associated with each selected metabolite at a genome-wide significant level (*p* < 5 × 10^−8^) from a genome-wide association study (GWAS).^(27)^ We then conducted linkage disequilibrium (LD) pruning at a threshold of 0.6 with minor allele frequency (MAF)>0.01 to exclude the genetic variants with high LD (https://ldlink.nci.nih.gov/?tab=snpclip). For each remaining genetic variant, we extracted the association results with FN-BMD and LS-BMD from the GWAS results reported by Estrada K. et al.^(28)^ Inverse-variance weighted (IVW) estimators then can be calculated to make causal inference between selected metabolites and FN-BMD or LS-BMD.^(29)^ We also calculated the weighted median estimator.^(30)^ We assessed the instrumental heterogeneity through the Q statistic and horizontal pleiotropy by Egger regression.^(31)^ IVW p-value less than 0.05 while both Q-statistic p-value and Egger regression intercept p-value greater than 0.05 was set as significance level. Significant outliers of instrumental variables can be detected by MR-PRESSO^(32)^. We conduct the power analysis according to Deng et al.^(33)^ We used the information about sample size of exposure and outcome, and proportion of exposure variance explained by the instrumental variables (r^2^) from the GWAS papers.^(27,28)^ We estimated the variance of exposure and outcome from our study data. Since we only selected a subset of variants which were genome-wide significant, we also included two other settings for r^2^: 80% of the given proportion from the GWAS paper; assumed that each genome-wide significant variant explained 0.01% of the variance. All analyses were conducted using R v3.5.3 program (https://cran.r-project.org/) except pathway enrichment analysis. Glmnet, pROC, PredictABEL and MendelianRandomization, four packages implemented in R, were used for the analysis of LASSO, risk prediction evaluation, and Mendelian randomization.

## Results

Conventional risk factors and osteoporosis-associated variables are shown in **Table 1**. In the discovery cohort, a total of 1,552 individuals were included in this analysis. The average age of participants was 55.4 years old. The proportion of women was 54.2% and 66% were post-menopausal. During a median follow-up period of 14 years (range 4-27) after the measurement of the plasma metabolites (i.e., baseline), 188 participants experienced at least one osteoporotic fracture. In the replication cohort, during a median of follow-up period of 10 years (range 0-22), 36 out of 634 participants experienced at least one osteoporotic fracture.

A total of 13 and 19 metabolites were selected at least 50 times out of 100 iterations to be associated with FN-BMD and LS-BMD in the discovery cohort, respectively (**Supplementary Tables 1 & 2**). The selected metabolites explained 17% variation of FN-BMD and 19% variation of LS-BMD, respectively. Overall, 27 metabolites were identified by their associations with either FN-BMD or LS-BMD (**Table 2**). Among them, five metabolites (glycine, leucine, pyridoxate, sphingomyelin [SM] C22:0, and xanthurenate) were associated with both FN-BMD and LS-BMD. One selected metabolite, fruc_gluc_galac, was a complex of fructose, glucose, and galactose, which cannot be grouped into any one category. Thus, there was no corresponding Human Metabolome Database (HMDB) ID for it. We also associated BMD with selected metabolites either altogether or individually in a model after adjusting all conventional risk factors. We refer interested readers to **Supplementary Table 3** for detail. One thing to mention is that only 18 out of 27 selected metabolites were available in the replication cohort. Therefore, all the results using replication cohort were based on those 18 available metabolites. Additionally, our secondary sex-stratified analysis in the discovery cohort identified more BMD-associated metabolites in female (15 and 10) than in male (1 and 2) associated with FN- and LS-BMD, respectively (**Supplementary Table 4**). Majority of them overlapped with above selected metabolites using the combined dataset.

**Table 2.** Twenty-seven metabolites were repeatedly selected ≥ 50 times in 100 iterations based on either FN-BMD or LS-BMD analysis.

| Name | Trait | Chemical class | Name | Trait | Chemical class |
| --- | --- | --- | --- | --- | --- |
| aconitate | FN BMD | Carboxylic acids and derivatives | PC C36:2 | FN BMD | Glycerophospholipids |
| ADP | LS BMD | Purine nucleotides | pyridoxate | FN & LS BMD | Pyridines and derivatives |
| creatine | LS BMD | Amino acids | serine | LS BMD | Amino acids |
| creatinine | LS BMD | Amino acids | serotonin | LS BMD | Indoles and derivatives |
| dimethylglycine | FN BMD | Amino acids | SM C16:1 | FN BMD | Organonitrogen compounds |
| fruc_gluc_galac | LS BMD | N.A. | SM C18:1 | LS BMD | Sphingolipids |
| glycine | FN & LS BMD | Amino acids | SM C22:0 | FN & LS BMD | Sphingolipids |
| hydroxyglutarate | FN BMD | Hydroxy acids and derivatives | sucrose | FN BMD | Carbohydrates and carbohydrate conjugates |
| hypoxanthine | LS BMD | Imidazopyrimidines | TAG C48:0 | LS BMD | Glycerolipids |
| leucine | FN & LS BMD | Amino acids | TAG C50:1 | LS BMD | Glycerolipids |
| LPC C18:1 | LS BMD | Glycerophospholipids | TAG C54:4 | LS BMD | Glycerolipids |
| LPE C18:2 | FN BMD | Glycerophospholipids | TAG C58:10 | LS BMD | Glycerolipids |
| pantothenate | FN BMD | Organooxygen compounds | xanthurenate | FN & LS BMD | Quinolines and derivatives |
| PC C36:1 | LS BMD | Glycerophospholipids |  |  |  |

BMD=bone mineral density. FN=femoral neck. LS=lumbar spine. SM=Sphingomyelin.
LPE=Lysophosphatidylethanolamine. PC=Phosphatidylcholine. LPC=Lysophosphatidylcholine.
TAG=Triacylglycerol. fruc\_gluc\_galac=Fructose+glucose+galactose. N.A.=not available

We present the adjusted R-squared results for the model comparisons in **Table 3**. Model 3 with conventional risk factors plus selected metabolites fit the data best with the highest adjusted R-squared for both discovery and replication cohorts (FN-BMD: overall adjusted R^2^ = 0.36 in the discovery cohort, and 0.39 in the replication cohort; LS-BMD: 0.28 for overall adjusted R^2^ in the discovery cohort, and 0.35 in the replication cohort) compared to model 1 with conventional risk factors alone (FN-BMD: 0.33 in the discovery cohort, and 0.36 in the replication cohort; LS-BMD: 0.23 in the discovery cohort, and 0.30 in the replication cohort) and model 2 with selected metabolites alone (FN-BMD: 0.16 in the discovery cohort, and; LS-BMD: 0.18 in the discovery cohort, and 0.13 in the replication cohort). Meanwhile, model 3 was significantly different from model 1 by F-test with overall p-value less than 0.001 for FN-BMD and LS-BMD in both discovery and replication cohorts. In the sensitivity analysis, additionally adjusting for diabetes status, drinking status, and calcium intake in the model 3, the model with clinical risk factors plus selected metabolites still fit the data best with the highest adjusted R^2^ and was significantly different from model 1 with p-value less than 0.001 for both FN-BMD and LS-BMD in the discovery cohort (**Supplementary Table 5**).

**Table 3.**
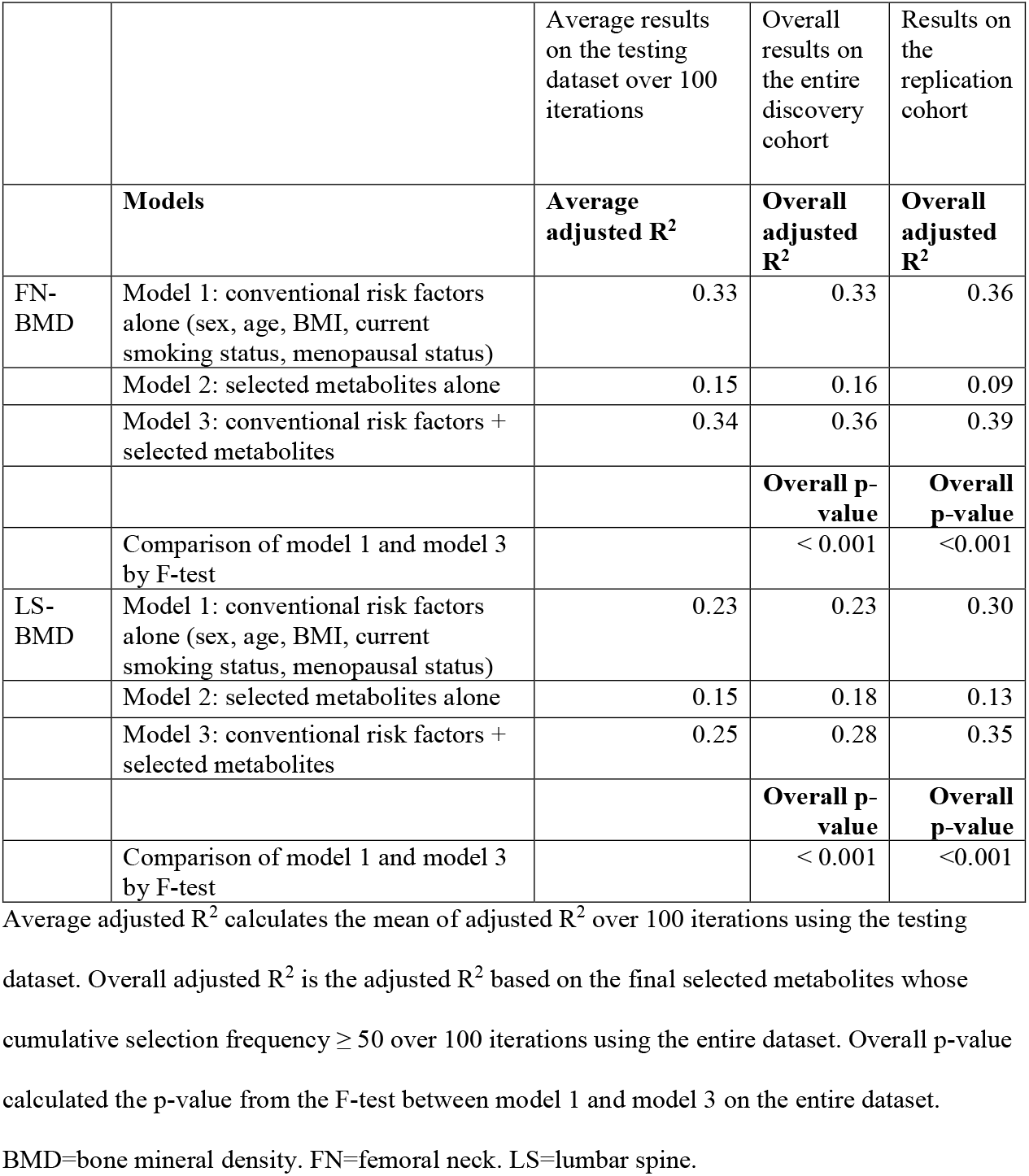
Average and overall results of model comparisons

In terms of fracture risk prediction, 27 selected metabolites significantly improved osteoporotic fracture risk prediction when added to conventional risk factors using the entire dataset (AUC = 0.74 [95% CI 0.70, 0.77] for model 2 with conventional risk factors + selected metabolites vs AUC = 0.70 [95% CI 0.66, 0.74] for model 1 with conventional risk factors alone, *p* = 0.001) (**Fig. 2A**). The difference in average predicted risks between the individuals with and without fracture increased by 3.1% in the model 2 (IDI=0.031, *p* < 0.001). Besides, when additionally accounting for FN-BMD or LS-BMD, the models with selected metabolites in addition to conventional risk factors still performed significantly better than the models with conventional risk factors alone (AUC = 0.75 [95% CI 0.72, 0.79] for model 2 with conventional risk factors + FN-BMD + selected metabolites vs AUC = 0.72 [95% CI 0.68, 0.76] for model 1 with conventional risk factors + FN-BMD, *p* = 0.002; AUC = 0.74 [95% CI 0.71, 0.78] for model 2 with conventional risk factors + LS-BMD + selected metabolites vs AUC = 0.71 [95% CI 0.67, 0.75] for model 1 with conventional risk factors + LS -BMD, *p* = 0.003) (**Fig. 2B & 2C**). The IDI=0.029 with *p* < 0.001 and IDI=0.032 with *p* < 0.001 for models additionally accounting for FN-BMD and LS-BMD, respectively. These results were replicated in the HKOS (**Table 4**).

**Table 4.** Model evaluation for fracture risk prediction

|  | Models | AUC | 95% CI of AUC | Delong's test p-value | IDI | IDI p-value | NRI | NRI p-value |
| --- | --- | --- | --- | --- | --- | --- | --- | --- |
| Discovery cohort | Model 1: conventional risk factors alone | 0.70 | 0.66, 0.74 | N.A. | N.A. | N.A. | N.A. | N.A. |
|  | Model 2: conventional risk factors + 27 selected metabolites | 0.74 | 0.70, 0.77 | 0.001 | 0.031 | <0.001 | 0.07 | 0.03 |
|  | Model 1+ FN-BMD | 0.72 | 0.68, 0.76 | N.A. | N.A. | N.A. | N.A. | N.A. |
|  | Model 2+ FN-BMD | 0.75 | 0.72, 0.79 | 0.002 | 0.029 | <0.001 | 0.08 | <0.001 |
|  | Model 1+ LS-BMD | 0.71 | 0.67, 0.75 | N.A. | N.A. | N.A. | N.A. | N.A. |
|  | Model 2+ LS-BMD | 0.74 | 0.71, 0.78 | 0.003 | 0.032 | <0.001 | 0.12 | <0.001 |
| Replication cohort | Model 1: conventional risk factors alone | 0.81 | 0.73, 0.89 | N.A. | N.A. | N.A. | N.A. | N.A. |
|  | Model 2: conventional risk factors + 18 selected metabolites | 0.87 | 0.81, 0.94 | 0.013 | 0.110 | <0.001 | 0.23 | 0.03 |
|  | Model 1+ FN-BMD | 0.87 | 0.80, 0.93 | N.A. | N.A. | N.A. | N.A. | N.A. |
|  | Model 2+ FN-BMD | 0.91 | 0.86, 0.96 | 0.015 | 0.079 | 0.003 | 0.09 | 0.16 |
|  | Model 1+ LS-BMD | 0.85 | 0.78, 0.92 | N.A. | N.A. | N.A. | N.A. | N.A. |
|  | Model 2+ LS-BMD | 0.89 | 0.83, 0.95 | 0.022 | 0.089 | 0.001 | 0.26 | 0.01 |

Model 1: conventional risk factors includes sex, age, BMI, current smoking status, menopausal status. Model 2: conventional risk factors which includes sex, age, BMI, current smoking status, menopausal status plus 27 or 18 selected metabolites. Participants were grouped into two categories, [0, 20%) and [20%, 1], based on their fracture risk for NRI. Delong's test, IDI, and
NRI were used to compare model 2 and model 1, and additionally accounting for FN-BMD or LS-BMD for both model 2 and model 1. AUC=Area under curve. IDI=Integrated discrimination improvement. NRI=Net reclassification index. CI=confidence interval. BMD=bone mineral density. FN=femoral neck. LS=lumbar spine. N.A.=not applicable

**Fig. 2.**
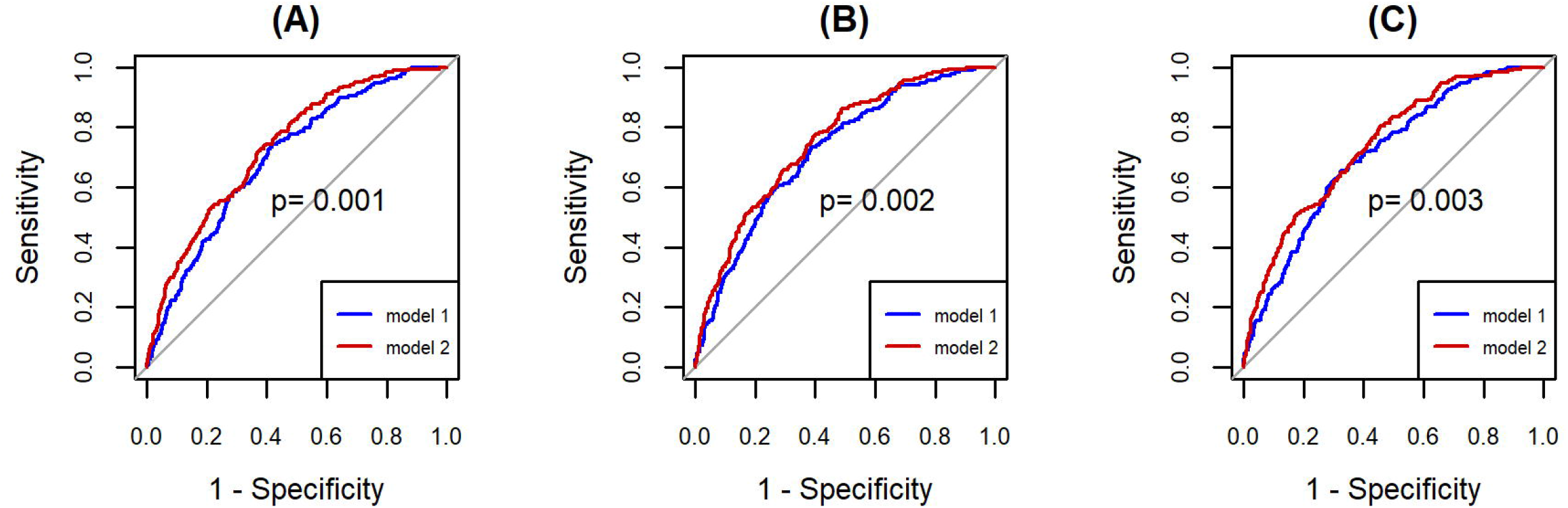
ROC curves of models for predicting osteoporotic fracture incidence with and without selected metabolites. (**A**) Blue line, model 1 with conventional risk factors alone: AUC = 0.70 (95% CI 0.66, 0.74); red line, model 2 with conventional risk factors + selected metabolites: AUC = 0.74 (95% CI 0.70, 0.77), with p-value=0.001. (**B**) Blue line, model 1 with conventional risk factors + FN-BMD: AUC = 0.72 (95% CI 0.68 0.76); red line, model 2 with conventional risk factors + FN-BMD + selected metabolites: AUC = 0.75 (95% CI 0.72, 0.79), with p-value=0.002. (**C**) Blue line, model 1 with conventional risk factors + LS-BMD: AUC = 0.71 (95% CI 0.67 0.75); red line, model 2 with conventional risk factors + LS-BMD + selected metabolites: AUC = 0.74 (95% CI 0.71, 0.78), with p-value=0.003. The p-value, calculated from DeLong’s test, less than 0.05 suggests the significant difference between two ROC curves. AUC=Area under curve. ROC=Receiver Operating Characteristic. BMD=bone mineral density. FN=femoral neck. LS=lumbar spine.

Reclassification table for subjects with and without a major osteoporotic fracture event during follow-up are summarized in **Table 5**. Take model 1 with conventional risk factor alone vs model 2 with conventional risk factor + selected metabolites as an example. For 23 individuals who experience a fracture event, model 2 with selected metabolites improved the classification (i.e. 23 individuals with events moved from the low-risk group [0, 20%) in model 1 to the high-risk group [20%, 1] in model 2). For 11 people with fracture event, the model 2 classification became worse (i.e. 11 individuals with events moved from the high-risk group [20%, 1] in model 1 to the low-risk group [0, 20%) in model 2). Similarly, for people who do not experience a fracture event, classification improved for 78 individuals and got worse for 70 individuals using the model 2 with selected metabolites. Overall, model 2 significantly improved classification of fracture risk with the NRI=0.07 (p-value=0.03). When additionally accounting for either FN-BMD or LS-BMD, model 2 with selected metabolites still significantly improved classification of fracture risk (FN-BMD: NRI=0.08, p-value < 0.001; LS-BMD: NRI=0.12, p-value < 0.001). These results were also replicated in the HKOS except for model 2 + FN-BMD vs model 1 + FN-BMD with p-value=0.16 (**Table 4**). In the sensitivity analysis with additional clinical risk factors in the model using the discovery cohort, incorporating selected metabolites still significantly improved osteoporotic fracture prediction (AUC=0.75 [95% CI 0.71, 0.78] for model 2 vs AUC=0.71 [95% CI 0.67, 0.75] for model 1, *p* = 0.002) although model 2 no longer significantly improved classification of fracture risk with the NRI=0.056 (p-value=0.14) (**Supplementary Table 6**).

**Table 5.** Reclassification table among people with and without a major osteoporotic fracture event during follow-up

| Models without selected metabolites | Models with selected metabolites |  |  |
| --- | --- | --- | --- |
| Model 1: Conventional risk factors alone | Model 2: Conventional risk factors + selected metabolites |  |  |
| Individuals who experience a major osteoporotic fracture event |  |  |  |
| Fracture risk | [0, 20%) | [20 %, 1] | Total |
| [0, 20%) | 102 | 23 | 125 |
| [20 %, 1] | 11 | 52 | 63 |
| Total | 113 | 75 | 188 |
| Individuals who do not experience a major osteoporotic fracture event |  |  |  |
| Fracture risk | [0, 20%) | [20 %, 1] | Total |
| [0, 20%) | 1092 | 70 | 1162 |
| [20 %, 1] | 78 | 124 | 202 |
| Total | 1170 | 194 | 1364 |
| Overall | NRI=0.07 | NRI p-value=0.03 |  |
| Model 1 + FN-BMD | Model 2 + FN-BMD |  |  |
| Individuals who experience a major osteoporotic fracture event |  |  |  |
| Fracture risk | [0, 20%) | [20 %, 1] | Total |
| [0, 20%) | 94 | 23 | 117 |
| [20 %, 1] | 9 | 62 | 71 |
| Total | 103 | 85 | 188 |
| Individuals who do not experience a major osteoporotic fracture event |  |  |  |
| Fracture risk | [0, 20%) | [20 %, 1] | Total |
| [0, 20%) | 1104 | 60 | 1164 |
| [20 %, 1] | 71 | 129 | 200 |
| Total | 1175 | 189 | 1364 |
| Overall | NRI=0.08 | NRI p-value < 0.001 |  |
| Model 1 + LS-BMD | Model 2 + LS-BMD |  |  |
| Individuals who experience a major osteoporotic fracture event |  |  |  |
| Fracture risk | [0, 20%) | [20 %, 1] | Total |
| [0, 20%) | 95 | 27 | 122 |
| [20 %, 1] | 8 | 58 | 66 |
| Total | 103 | 85 | 188 |
| Individuals who do not experience a major osteoporotic fracture event |  |  |  |
| Fracture risk | [0, 20%) | [20 %, 1] | Total |
| [0, 20%) | 1103 | 57 | 1160 |
| [20 %, 1] | 76 | 128 | 204 |
| Total | 1179 | 185 | 1364 |
| Overall | NRI=0.12 | NRI p-value < 0.001 |  |

Model 1: conventional risk factors includes sex, age, BMI, current smoking status, menopausal status. Model 2: conventional risk factors which includes sex, age, BMI, current smoking status, menopausal status plus 27 selected metabolites. Logistic regression was fitted to get the predicted probability of fracture as fracture risk here for all models. Participants were grouped into two categories, [0, 20%) and [20%, 1], based on their fracture risk. NRI=Net reclassification index. CI=confidence interval. BMD=bone mineral density. FN=femoral neck. LS=lumbar spine.

Based on the selected 27 metabolites, the glycine, serine, and threonine metabolism pathway including four identified metabolites (creatine, dimethylglycine, glycine, and serine) was significantly enriched with an FDR adjusted p-value=0.028. We also performed two-sample Mendelian randomization to investigate the causal effect of these selected metabolites on BMD. We focused on six selected metabolites (leucine, glycine, creatinine, phosphatidylcholine [PC], sphingomyelin [SM], and triacylglycerol [TAG]) since the genetic association data for the other selected metabolites were not available in the GWAS ^(27)^. Three selected metabolites (glycine, PC, and TAG) had a causally negative association with FN-BMD and two (PC and TAG) had a causally negative association with LS-BMD at a significance level of 0.05 based on IVW p-value while heterogeneity and horizontal pleiotropy were not observed (i.e. neither p-value of Q statistic nor p-value of Egger regression intercept was less than 0.05) (**Table 6**). In particular, for every 1 standard deviation increase in glycine, the value of FN-BMD decreases 0.033 in g/cm^2^. Similarly, LS-BMD decreased by 0.136 g/cm^2^ for each standard deviation increase in PC. Note that PC with FN-BMD as outcome and TAG with either FN- or LS-BMD as outcome are no longer significant based on the weighted median method (**Table 6**). We did not observe any significant outliers through MR-PRESSO. All the related genetic variants are available in **Supplementary Table 7**. The power analysis under type I error (*α* = 0.05) demonstrates insufficient power (estimated power < 0.4) to detect the significant association between leucine and creatinine with either FN- or LS-BMD, glycine with LS-BMD, and SM with FN-BMD for all three settings of r^2^ (**Supplementary Table 8**). Therefore, those insignificant associations between selected metabolites and BMD could be due to limited statistical power.

**Table 6.** Two-sample Mendelian randomization results for FN-BMD and LS-BMD

| Exposure (Metabolite) | Outcome (BMD) | IVW beta | IVW p-value | Weighted median beta | Weighted median p-value | Q statistic p-value | Egger regression intercept p-value |
| --- | --- | --- | --- | --- | --- | --- | --- |
| leucine | FN BMD | -0.028 | 0.63 | -0.057 | 0.34 | 0.22 | 0.75 |
|  | LS BMD | 0.071 | 0.19 | 0.074 | 0.24 | 0.35 | 0.72 |
| **glycine** | FN BMD | -0.033 | $2.51 \times 10^{-5}$ | -0.040 | $6.82 \times 10^{-4}$ | 0.98 | 0.87 |
|  | LS BMD | -0.013 | 0.14 | -0.017 | 0.16 | 0.89 | 0.71 |
| creatinine | FN BMD | 0.074 | 0.39 | 0.169 | 0.03 | 0.01 | $1.08 \times 10^{-4}$ |
| | LS BMD | 0.146 | 0.02 | 0.164 | 0.04 | 0.16 | $3.90 \times 10^{-3}$ |
| **PC** | FN BMD | -0.066 | $6.89 \times 10^{-3}$ | -0.043 | 0.19 | 0.84 | 0.05 |
| | LS BMD | -0.136 | $1.84 \times 10^{-7}$ | -0.106 | $2.96 \times 10^{-3}$ | 0.57 | 0.30 |
| SM | FN BMD | -0.017 | 0.75 | -0.011 | 0.84 | 0.06 | 0.21 |
| | LS BMD | -0.145 | 0.01 | -0.164 | $7.41 \times 10^{-3}$ | 0.05 | 0.03 |
| **TAG** | FN BMD | -0.065 | $7.46 \times 10^{-3}$ | -0.033 | 0.35 | 0.78 | 0.59 |
|  | LS BMD | -0.070 | 0.02 | -0.039 | 0.35 | 0.18 | 0.54 |

Genetic information of above six metabolites (exposure) and two outcomes, FN-BMD and LS-
BMD, are available from the summary statistics of GWAS.<sup>23,24</sup> IVW (inverse variance weighted)
beta estimates the causal effect of each selected metabolite on BMD, either FN-BMD or LS-
BMD. Q statistic p-value less than 0.05 indicates the instrumental heterogeneity for each selected
metabolite. Egger regression intercept p-value less than 0.05 implicates the horizontal pleiotropy
for the instrumental variants of each selected metabolite. IVW p-value less than 0.05 while both
Q-statistics p-value and Egger regression intercept p-value greater than 0.05 was set as the
significance level. Bold ones are the metabolites with significant causal effect on either FN-
BMD or LS-BMD. BMD=bone mineral density. FN=femoral neck. LS=lumbar spine.

## Discussion

We performed an association study among FHS Offspring participants relating plasma metabolites to both FN-BMD and LS-BMD, and we assessed the use of metabolites for predicting the risk of major osteoporotic fractures over a follow-up period exceeding two decades. We identified 27 metabolites that were associated with FN-BMD or LS-BMD. Incorporating these metabolites into a prediction model for fractures significantly improved the prediction of osteoporotic fracture risk beyond the conventional clinical risk factors including sex, age, BMI, current smoking status, and menopausal status. These results were replicated in an independent study and showed the potential of metabolites in the prediction of fracture. Additionally, the glycine, serine, and threonine metabolism pathway (including four identified metabolites: creatine, dimethylglycine, glycine, and serine) was significantly enriched. Among six selected metabolites, using Mendelian randomization, three (glycine, PC, and TAG) were found to be causally negatively associated with FN-BMD. Two of these metabolites (PC and TAG) were causally negatively associated with LS-BMD. It suggested that those metabolites may contribute to alterations in BMD and may relate to osteoporosis pathogenesis.

A major clinical implication of our study is that incorporating selected metabolites can help improve osteoporotic fracture risk prediction beyond conventional clinical risk factors. Previous studies have only shown that incorporating metabolites can improve the classification of people with different levels of BMD, but not directly on osteoporotic fracture risk prediction.^(9-11)^ Our study additionally demonstrated that lipid profiles (11 out of 27 selected metabolites are lipid-related) and amino acids (six out of 27 selected metabolites belong to amino acids) might involve in bone metabolism, which was consistent with previous studies (**Table 2**).^(11,34)^ We not only confirmed previous reports of associations between two selected plasma metabolite (dimethylglycine, creatinine) and BMD,^(9,13)^ we also identified many novel metabolites compared to other studies. One possible reason could be that different types of metabolites were measured across different studies without a uniform standard, which suggests that a harmonized platform for metabolomic analysis is needed for future studies.

The glycine, serine and threonine metabolism pathway was enriched based on the selected metabolites in our study, which was consistent with a recent association analysis finding reported by Zhao Q. and colleagues.^(10)^ While there is no known direct mechanism to explain our pathway enrichment results related to osteoporosis, three (creatine, dimethylglycine, and glycine) out of the four selected metabolites involved in this pathway have been reported to be associated with bone health in various ways. The creatine/phosphorylcreatine system is involved in bioenergetic processes, especially in tissues with high metabolic demand such as skeletal muscle and bone.^(35)^ A recent review suggested that creatine may affect the bone remodeling process and have beneficial effects on lean mass and muscle for older individuals.^(35)^ Dimethylglycine belongs to the choline oxidation pathway and is demethylated in the mitochondria, leading to the subsequent formation of glycine, which is a non-essential amino acid.^(36)^ Low plasma dimethylglycine was found to be associated with low BMD and an increased risk of hip fractures by Øyen J et al.^(37)^ Men with idiopathic osteoporosis had a higher plasma glycine.^(38)^ Our Mendelian randomization results (**Table 6**) also supported that glycine was causally negatively associated with FN-BMD. In other words, higher plasma glycine may result in lower FN-BMD and osteoporosis. In addition, our Mendelian randomization analysis found that PC and TAG may causally result in lower FN- and LS-BMD (**Table 6**). PC and TAG are both lipids. Several studies suggest that lipid profiles are associated with BMD and might be useful for osteoporosis prediction.^(11,12)^ One study reported that higher concentrations of phosphatidylcholine docosahexaenoic acid (PC DHA) was associated with loss of FN-BMD over 4 years in women, where PC is the backbone of PC DHA.^(39)^ Overall, though our identified metabolites may be of potential clinical value for the early detection of osteoporosis, further experimental validation and clinical replication are required. We anticipate that based on studies like ours and those of others, these data will guide future hypothesis-driven analyses for better understanding of the relationship between metabolites and skeletal traits.

Due to the difficulties in harmonizing metabolite platforms, one potential limitation of this study was the external validation. While our results were in general replicated, not all of our selected metabolites were available in our replication cohort. A second limitation of our investigation was that only 209 metabolites were measured. As technology advances, the number of identifiable metabolites increases. We believe that including more metabolites in a future association study is meaningful to get a more comprehensive understanding of the relationship between metabolites and osteoporosis. Third, BMD were measured two to ten years later after metabolites measured and the storage time for metabolites were about 15-20 years in our discovery and require more validations even though we had replicated our results in an independent study. Lastly, we did not conduct the analysis to account for time-varying covariates. Such analysis also be of interest because the metabolite’s measurements may change over time. This would be a potential future direction with newly generated data.

In conclusion, we identified 27 metabolites that were associated BMD and helpful in osteoporotic fracture risk prediction. The glycine, serine and threonine metabolism pathway was significantly enriched based on our selected metabolites. Three selected metabolites (glycine, PC, and TAG) had causally negative associations with BMD. These identified metabolites may have a role in the early stages of osteoporosis and may provide novel insights into the mechanisms underlying this disease. Our findings raise the possibility of using metabolite profiling to improve the prediction of osteoporotic fracture if confirmed by other studies.

## Data Availability

Framingham Cohort data is available in dbGaP.

## Supplemental Data

Supplemental Data include eight tables.

## Disclosures

The authors declare no conflict of interest.

## Acknowledgements

This work was partly supported by NHLBI/NIH Framingham Heart Study contract numbers: NO1-HC-25195 (RSV), HHSN268201500001I (RSV), & 75N92019D00031 (RSV), NIDDK/NIH R01 DK081572 (RSV), NIAMS/NIH R01 AR041398 (DPK, CTL), NIAM/NIH R01 AR072199 (YHH, HX, CTL), Doris Duke Charitable Foundation (XZ, MTL, CTL), the Evans Medical Foundation and the Jay and Louis Coffman Endowment from the Department of Medicine, Boston University School of Medicine (RSV).The replication study is supported by Early Career Scheme (27110416) funded by the Research Grants Council, HKSAR, China.

Authors’ roles: CTL conceived of the idea and supervised the work. XZ and CTL designed the model and framework. XZ and GHYL carried out the analysis. XZ prepared the original draft. All authors (XZ, HX, GHYL, MTL, CLC, RSV, YHH, DPK, CTL) discussed the results, provided critical feedbacks and helped shape the manuscript.

## Notes

### Competing Interest Statement

The authors have declared no competing interest.

### Author Declarations

All participants provided written informed consent and the study protocol was approved by the Hebrew SeniorLife and Boston University Medical Campus institutional review boards.

