## Supplemental material for "Metabolomics insights into osteoporosis through association with bone mineral density"

**Supplementary Table 1.** 13 metabolites were repeatedly selected ≥ 50 times in 100 iterations based on FN BMD analysis in the discovery cohort

| **Name** | **HMDB-ID** | **Selected times (out of 100 iterations)** |
| --- | --- | --- |
| leucine | HMDB0000687 | 97 |
| xanthurenate | HMDB0000881 | 83 |
| aconitate | HMDB0000072 | 80 |
| pyridoxate | HMDB0000017 | 76 |
| SM C22:0 | HMDB0012091 | 73 |
| sucrose | HMDB0000258 | 70 |
| PC C36:2 | HMDB0008071 | 65 |
| glycine | HMDB0000123 | 59 |
| SM C16:1 | HMDB0029216 | 57 |
| LPE C18:2 | HMDB0011507 | 53 |
| hydroxyglutarate | HMDB0000428 | 53 |
| pantothenate | HMDB0000210 | 52 |
| dimethylglycine | HMDB0000092 | 50 |

BMD=bone mineral density. FN=femoral neck. SM=Sphingomyelin. PC=Phosphatidylcholine. LPE=Lysophosphatidylethanolamine.

**Supplementary Table 2** 19 metabolites were repeatedly selected ≥ 50 times in 100 iterations based on LS BMD analysis in the discovery cohort

| **Name** | **HMDB-ID** | **Selected times (out of 100 iterations)** |
| --- | --- | --- |
| pyridoxate | HMDB0000017 | 98 |
| fruc_gluc_galac | NA | 92 |
| SM C18:1 | HMDB0010169 | 90 |
| TAG C50:1 | HMDB0005360 | 86 |
| serotonin | HMDB0000259 | 83 |
| leucine | HMDB0000687 | 82 |
| serine | HMDB0000187 | 79 |
| xanthurenate | HMDB0000881 | 67 |
| glycine | HMDB0000123 | 67 |
| creatinine | HMDB0000562 | 67 |
| hypoxanthine | HMDB0000157 | 63 |
| adp | HMDB0001341 | 61 |
| TAG C54:4 | HMDB0005455 | 58 |
| creatine | HMDB0000064 | 56 |
| LPC C18:1 | HMDB0010385 | 53 |
| PC C36:1 | HMDB0008038 | 53 |
| TAG C58:10 | HMDB0005476 | 53 |
| SM C22:0 | HMDB0012091 | 51 |
| TAG C48:0 | HMDB0005356 | 50 |

BMD=bone mineral density. LS=lumbar spine. fruc_gluc_galac=Fructose+glucose+galactose.SM=Sphingomyelin. TAG=Triacylglycerol. LPC=Lysophosphatidylcholine. HMDB=Human Metabolome Database. PC=Phosphatidylcholine. NA=not applicable.

**Supplementary Table 3.** Association results for FN-BMD and LS-BMD with full model (conventional risk factors + all selected metabolites) and individual model (conventional risk factors + each selected metabolite) in the discovery and replication cohorts

| **Discovery cohort** | | | | | **Replication cohort** | | | | |
| --- | --- | --- | --- | --- | --- | --- | --- | --- | --- |
| **FN-BMD** | **Full model** | | **Individual model** | | **FN-BMD** | **Full model** | | **Individual model** | |
| **Metabolite  Name** | **Estimate (SE)** | **P-value** | **Estimate (SE)** | **P-value** | **Metabolite  Name** | **Estimate (SE)** | **P-value** | **Estimate (SE)** | **P-value** |
| aconitate | -0.0092 (0.003) | 0.0061 | -0.0105 (0.003) | 0.0014 | aconitate | -0.0076(0.006) | 0.216 | -0.0087 (0.006) | 0.120 |
| dimethylglycine | -0.0098 (0.003) | 0.0032 | -0.0092 (0.003) | 0.0053 | dimethylglycine | 0.013 (0.006) | 0.020 | 0.015 (0.006) | 0.0062 |
| glycine | -0.0053 (0.003) | 0.0985 | -0.0078 (0.003) | 0.0144 | glycine | 0.003 (0.009) | 0.732 | -0.011 (0.005) | 0.048 |
| hydroxyglutarate | -0.0049 (0.003) | 0.1184 | -0.0074 (0.003) | 0.0172 | hydroxyglutarate | -0.018 (0.006) | 0.001 | -0.019 (0.005) | 0.0005 |
| leucine | 0.0121 (0.004) | 0.0015 | 0.0112  (0.004) | 0.0026 | leucine | 0.0006 (0.005) | 0.009 | -0.011 (0.006) | 0.046 |
| LPE C18:2 | -0.0053 (0.004) | 0.1679 | -0.0115 (0.003) | 0.0002 | LPE C18:2 | 0.010 (0.006) | 0.084 | 0.009 (0.005) | 0.093 |
| pantothenate | 0.0068 (0.004) | 0.0960 | 0.0091  (0.003) | 0.0037 | pantothenate | 0.013 (0.006) | 0.044 | 0.014 (0.005) | 0.013 |
| PC C36:2 | -0.0032 (0.004) | 0.4358 | -0.0110 (0.003) | 0.0004 | N.A. | N.A. | N.A. | N.A. | N.A. |
| pyridoxate | 0.0048 (0.004) | 0.2439 | 0.0098  (0.003) | 0.0018 | pyridoxate | -0.0011(0.006) | 0.858 | 0.0061 (0.005) | 0.263 |
| SM C16:1 | -0.0022 (0.004) | 0.5958 | -0.0119 (0.003) | 0.0004 | N.A. | N.A. | N.A. | N.A. | N.A. |
| SM C22:0 | -0.0070 (0.004) | 0.0614 | -0.0109 (0.003) | 0.0004 | SM C22:0 | -0.006 (0.006) | 0.277 | -0.0027 (0.006) | 0.643 |
| sucrose | -0.0059 (0.003) | 0.0528 | -0.0074 (0.003) | 0.0163 | sucrose | -0.0058 (0.005) | 0.294 | -0.0082 (0.005) | 0.127 |
| xanthurenate | 0.0096 (0.003) | 0.0025 | 0.0101  (0.003) | 0.0013 | xanthurenate | 0.013 (0.006) | 0.018 | 0.015 (0.005) | 0.0053 |
| **LS-BMD** | **Full model** | | **Individual model** | | **LS-BMD** | **Full model** | | **Individual model** | |
| **Metabolite  Name** | **Estimate (SE)** | **P-value** | **Estimate (SE)** | **P-value** | **Metabolite  Name** | **Estimate (SE)** | **P-value** | **Estimate (SE)** | **P-value** |
| adp | -0.0056 (0.0052) | 0.2828 | -0.0132 (0.0047) | 0.0051 | N.A. | N.A. | N.A. | N.A. | N.A. |
| creatine | -0.0076 (0.0054) | 0.1610 | -0.0012 (0.0053) | 0.8202 | creatine | -0.013 (0.007) | 0.073 | -0.016 (0.007) | 0.025 |
| creatinine | 0.0077  (0.0055) | 0.1654 | 0.0082 (0.0053) | 0.1208 | creatinine | 0.038 (0.009) | 3.78E-5 | 0.044 (0.008) | 2.30E-7 |
| fruc_gluc_galac | 0.0159  (0.0050) | 0.0017 | 0.0177 (0.0050) | 0.0004 | N.A. | N.A. | N.A. | N.A. | N.A. |
| glycine | -0.0050 (0.0055) | 0.3595 | -0.0143 (0.0049) | 0.0036 | glycine | -0.0041 (0.014) | 0.766 | -0.016 (0.007) | 0.023 |
| hypoxanthine | 0.0114  (0.0047) | 0.0159 | 0.0096 (0.0047) | 0.0416 | hypoxanthine | -0.0044(0.007) | 0.500 | -0.012 (0.007) | 0.091 |
| leucine | 0.0128  (0.0063) | 0.0414 | 0.0148 (0.0057) | 0.0092 | leucine | 0.030 (0.017) | 0.077 | -0.011 (0.007) | 0.108 |
| LPC C18:1 | -0.0082 (0.0059) | 0.1657 | -0.0231 (0.0048) | 1.85E-5 | N.A. | N.A. | N.A. | N.A. | N.A. |
| PC C36:1 | -0.0133 (0.0063) | 0.0343 | -0.0081 (0.0048) | 0.0888 | PC C36:1 | -0.019 (0.007) | 0.0094 | -0.014 (0.007) | 0.043 |
| pyridoxate | 0.0186  (0.0049) | 0.0002 | 0.0187 (0.0048) | 0.0001 | pyridoxate | 0.0036 (0.007) | 0.605 | 0.012 (0.007) | 0.099 |
| serine | -0.0064 (0.0057) | 0.2568 | -0.0160 (0.0048) | 0.0008 | serine | -0.039 (0.021) | 0.070 | -0.018 (0.007) | 0.0076 |
| serotonin | -0.0123 (0.0053) | 0.0195 | -0.0124 (0.0047) | 0.0084 | serotonin | 0.0007 (0.007) | 0.922 | 0.0085 (0.007) | 0.210 |
| SM C18:1 | -0.0072 (0.0057) | 0.2095 | -0.0214 (0.0050) | 1.63E-5 | SM C18:1 | 0.0035 (0.008) | 0.664 | -0.0078 (0.007) | 0.265 |
| SM C22:0 | -0.0052 (0.0052) | 0.3168 | -0.0138 (0.0047) | 0.0035 | SM C22:0 | -0.014 (0.008) | 0.068 | -0.014 (0.007) | 0.061 |
| TAG C48:0 | 0.0055  (0.0066) | 0.4007 | 0.0164 (0.0050) | 0.0010 | N.A. | N.A. | N.A. | N.A. | N.A. |
| TAG C50:1 | 0.0152  (0.0050) | 0.0025 | 0.0143 (0.0047) | 0.0024 | N.A. | N.A. | N.A. | N.A. | N.A. |
| TAG C54:4 | -0.0095 (0.0052) | 0.0672 | -0.0157 (0.0047) | 0.0009 | N.A. | N.A. | N.A. | N.A. | N.A. |
| TAG C58:10 | 0.0119  (0.0050) | 0.0181 | 0.0111 (0.0048) | 0.0206 | N.A. | N.A. | N.A. | N.A. | N.A. |
| xanthurenate | 0.0066  (0.0049) | 0.1796 | 0.0142 (0.0048) | 0.0032 | xanthurenate | 0.020 (0.007) | 0.0037 | 0.025 (0.007) | 0.0003 |

SE=standard error. BMD=bone mineral density. FN=femoral neck. LS=lumbar spine. SM=Sphingomyelin. LPE=Lysophosphatidylethanolamine. PC=Phosphatidylcholine. LPC=Lysophosphatidylcholine. TAG=Triacylglycerol. fruc_gluc_galac=Fructose+glucose+galactose.N.A.=not available in the replicated study.

**Supplementary Table 4.** Sex-stratified analysis to select metabolites by FN-BMD or LS-BMD with frequency ≥ 50 over 100 iterations in the discovery cohort

| **FN-BMD** | | | **LS-BMD** | | |
| --- | --- | --- | --- | --- | --- |
| **Female** | | | **Female** | | |
| **Name** | **HMDB-ID** | **Selected times (out of 100 iterations)** | **Name** | **HMDB-ID** | **Selected times (out of 100 iterations)** |
| **SM C22:0** | HMDB0012091 | 88 | **LPC C18:1** | HMDB0010385 | 88 |
| **LPC C18:1** | HMDB0010385 | 86 | **SM C18:1** | HMDB0010169 | 83 |
| cystathionine | HMDB0000099 | 86 | **TAG C58:10** | HMDB0005476 | 63 |
| **sucrose** | HMDB0000258 | 81 | **pyridoxate** | HMDB0000017 | 60 |
| malate | HMDB0000156 | 77 | **xanthurenate** | HMDB0000881 | 58 |
| **pantothenate** | HMDB0000210 | 77 | deoxycholates | HMDB0000626 | 58 |
| CE C18:3 | HMDB0010369 | 70 | orotate | HMDB0000226 | 56 |
| CE C18:1 | HMDB0000918 | 67 | kynurenine | HMDB0000684 | 54 |
| ornithine | HMDB0000214 | 67 | gdp | HMDB0001201 | 53 |
| SDMA | HMDB0003334 | 59 | betaine | HMDB0000043 | 52 |
| **TAG C58:10** | HMDB0005476 | 57 |  |  |  |
| lactose | HMDB0000186 | 55 |  |  |  |
| **xanthurenate** | HMDB0000881 | 55 |  |  |  |
| **leucine** | HMDB0000687 | 54 |  |  |  |
| **LPE C18:2** | HMDB0011507 | 52 |  |  |  |
| **Male** | | | **Male** | | |
| isocitrate | HMDB0000193 | 56 | lactate | HMDB0144295 | 56 |
|  |  |  | **hypoxanthine** | HMDB0000157 | 51 |

Bold metabolites are those overlapped with the selected metabolites using the combined dataset either by FN-BMD or LS-BMD. BMD=bone mineral density. FN=femoral neck. LS=lumbar spine. SM=Sphingomyelin. LPE=Lysophosphatidylethanolamine. CE=Cholesterol ester. LPC=Lysophosphatidylcholine. TAG=Triacylglycerol

**Supplementary Table 5.** Sensitivity analysis incorporating additional covariates in model comparison when BMD as outcome n=1350 in the discovery cohort

|  | **Models** | **Adjusted R^2^** |
| --- | --- | --- |
| FN-BMD | Model 1: conventional risk factors alone (sex, age, BMI, current smoking status, menopausal status, diabetes status, drinking status, calcium intake) | 0.32 |
|  | Model 2: selected metabolites alone | 0.16 |
|  | Model 3: conventional risk factors above + selected metabolites | 0.35 |
|  |  | **F-test p-value** |
|  | Comparison of model 1 and model 3 by F-test | < 0.001 |
|  |  | **Adjusted R^2^** |
| LS-BMD | Model 1: conventional risk factors alone (sex, age, BMI, current smoking status, menopausal status, diabetes status, drinking status, calcium intake) | 0.24 |
|  | Model 2: selected metabolites alone | 0.18 |
|  | Full model: conventional risk factors above+ selected metabolites | 0.28 |
|  |  | **F-test p-value** |
|  | Comparison of model 1 and model 3 by F-test | < 0.001 |

BMD=bone mineral density. FN=femoral neck. LS=lumbar spine.

**Supplementary Table 6.** Sensitivity analysis incorporating additional covariates in model comparison when fracture as outcome n=1350 in the discovery cohort

|  | **AUC** | **95% CI of AUC** | **Delong’s test p-value** | **IDI** | **IDI p-value** | **NRI** | **NRI p-value** |
| --- | --- | --- | --- | --- | --- | --- | --- |
| Model 1: conventional risk factors (sex, age, BMI, current smoking status, menopausal status, diabetes status, drinking status, calcium intake) | 0.71 | 0.67, 0.75 | N.A. | N.A. | N.A. | N.A. | N.A. |
| Model 2: conventional risk factors above + 27 selected metabolites | 0.75 | 0.71, 0.78 | 0.002 | 0.029 | < 0.001 | 0.056 | 0.14 |
| Model 1+ FN-BMD | 0.73 | 0.69 0.77 | N.A. | N.A. | N.A. | N.A. | N.A. |
| Model 2+ FN-BMD | 0.76 | 0.73, 0.80 | 0.002 | 0.028 | < 0.001 | 0.095 | 0.003 |
| Model 1+ LS-BMD | 0.72 | 0.68, 0.76 | N.A. | N.A. | N.A. | N.A. | N.A. |
| Model 2+ LS-BMD | 0.75 | 0.72, 0.79 | 0.004 | 0.03 | < 0.001 | 0.116 | < 0.001 |

BMD=bone mineral density. FN=femoral neck. LS=lumbar spine. N.A.=not applicable. AUC=area under curve. NRI=net reclassification index. IDI=integrated discrimination improvement.

**Supplementary Table 7.** Characteristics of genetic variants considered for use in Mendelian randomization analysis of the effect of exposure and outcomes (FN-BMD, LS-BMD)

| **Exposure** | **rsid** | **Chromosome** | **Position** | **EA** | **EAF** | **Exposure beta** | **Exposure p-value** | **FN-BMD beta** | **FN-BMD p-value** | **LS-BMD beta** | **LS-BMD p-value** |
| --- | --- | --- | --- | --- | --- | --- | --- | --- | --- | --- | --- |
| Leucine | rs17789027 | 4 | 89181841 | G | 0.384329 | 0.108594 | 6.16E-30 | -0.0106 | 0.2259 | -0.0026 | 0.7783 |
| Leucine | rs7678928 | 4 | 89222827 | T | 0.425537 | 0.100105 | 2.15E-26 | 0.0018 | 0.8311 | 0.0165 | 0.06422 |
| Leucine | rs893970 | 4 | 89236735 | C | 0.446964 | 0.078543 | 1.2E-14 | -0.009 | 0.298 | 0.005 | 0.5839 |
| Leucine | rs12325419 | 16 | 70368909 | G | 0.879779 | 0.082062 | 4.55E-08 | 0.0178 | 0.1422 | -0.0148 | 0.2452 |
| Glycine | rs10167637 | 2 | 211246007 | C | 0.128927 | 0.09289 | 2.68E-09 | -0.0043 | 0.7403 | 0.0013 | 0.9298 |
| Glycine | rs10195020 | 2 | 211657226 | T | 0.491955 | 0.097562 | 1.85E-18 | -0.002 | 0.8126 | 0.0029 | 0.745 |
| Glycine | rs10205894 | 2 | 211657098 | G | 0.573074 | 0.073947 | 5.47E-12 | -0.0029 | 0.7278 | 0.0057 | 0.5176 |
| Glycine | rs10804186 | 2 | 211495110 | T | 0.457274 | 0.061514 | 1.12E-08 | 0.0078 | 0.3492 | 0.0104 | 0.2434 |
| Glycine | rs10804193 | 2 | 212196284 | G | 0.34016 | 0.075728 | 2E-10 | -0.0047 | 0.6085 | 0.0237 | 0.01522 |
| Glycine | rs10932348 | 2 | 211538455 | A | 0.882486 | 0.129598 | 1.85E-12 | -0.0651 | 0.02324 | -0.1008 | 0.01068 |
| Glycine | rs11675189 | 2 | 211722595 | C | 0.883056 | 0.115294 | 1.04E-12 | -0.0143 | 0.3497 | -0.0238 | 0.1454 |
| Glycine | rs11689581 | 2 | 212598084 | C | 0.222657 | 0.078296 | 7.7E-10 | -0.0055 | 0.6092 | -0.0119 | 0.2992 |
| Glycine | rs11895699 | 2 | 211865593 | A | 0.858947 | 0.112936 | 3.35E-13 | -0.0137 | 0.2419 | -0.0121 | 0.334 |
| Glycine | rs12464940 | 2 | 211682987 | T | 0.783709 | 0.109138 | 1.02E-15 | -0.0141 | 0.1725 | -0.0175 | 0.1129 |
| Glycine | rs12478788 | 2 | 211681243 | A | 0.369198 | 0.227241 | 9.67E-87 | -0.0059 | 0.5104 | -0.0023 | 0.8097 |
| Glycine | rs12694212 | 2 | 211695915 | A | 0.818955 | 0.083258 | 9.69E-10 | -0.0143 | 0.2093 | -0.023 | 0.05953 |
| Glycine | rs13003126 | 2 | 211711928 | C | 0.871109 | 0.122611 | 4.95E-15 | -0.0107 | 0.4056 | -0.0077 | 0.5768 |
| Glycine | rs13010668 | 2 | 211470493 | G | 0.163462 | 0.078735 | 2.88E-08 | -0.006 | 0.5879 | 0.0018 | 0.8786 |
| Glycine | rs13011429 | 2 | 211595331 | G | 0.869556 | 0.18599 | 1.63E-29 | -0.0186 | 0.1616 | 0.017 | 0.2389 |
| Glycine | rs13018117 | 2 | 212705380 | A | 0.079377 | 0.157989 | 2.88E-14 | 0.046 | 0.3811 | -0.1233 | 0.05727 |
| Glycine | rs13019409 | 2 | 211701811 | G | 0.38252 | 0.193259 | 8.26E-64 | 0.0013 | 0.8847 | 0.0004 | 0.9662 |
| Glycine | rs13022335 | 2 | 211514198 | C | 0.719248 | 0.068525 | 1.63E-08 | -0.0142 | 0.1954 | 0.001 | 0.933 |
| Glycine | rs13026255 | 2 | 212596915 | C | 0.367795 | 0.063884 | 2.77E-08 | 0.0036 | 0.6734 | 0.0075 | 0.4058 |
| Glycine | rs13027153 | 2 | 211675576 | A | 0.8517 | 0.143606 | 1.86E-21 | -0.0086 | 0.5175 | 0.0082 | 0.5756 |
| Glycine | rs13386028 | 2 | 211547400 | C | 0.573389 | 0.279638 | 4.45E-156 | -0.015 | 0.0718 | 0.0081 | 0.3636 |
| Glycine | rs13394185 | 2 | 211655017 | C | 0.915413 | 0.107858 | 1.12E-08 | 0.023 | 0.752 | -0.0089 | 0.9074 |
| Glycine | rs13395652 | 2 | 212303506 | A | 0.687668 | 0.066159 | 4.31E-08 | -0.0055 | 0.5347 | -0.0121 | 0.2053 |
| Glycine | rs13399140 | 2 | 211431074 | C | 0.953574 | 0.154477 | 2.16E-09 | 0.0059 | 0.8086 | 0.0058 | 0.8242 |
| Glycine | rs16844788 | 2 | 211540863 | T | 0.883664 | 0.191866 | 1.67E-28 | 0.0069 | 0.6379 | 0.0119 | 0.4548 |
| Glycine | rs17678015 | 2 | 211739710 | C | 0.304609 | 0.269425 | 4.39E-124 | -0.0038 | 0.7015 | 0.0028 | 0.7959 |
| Glycine | rs17775871 | 2 | 211548674 | G | 0.089676 | 0.250821 | 1.56E-40 | -0.0056 | 0.6262 | 0.0034 | 0.7878 |
| Glycine | rs17792034 | 2 | 211751804 | A | 0.876123 | 0.116378 | 1.24E-13 | -0.0184 | 0.2127 | -0.0159 | 0.3115 |
| Glycine | rs1821657 | 2 | 212168303 | T | 0.224762 | 0.074271 | 3.3E-09 | 0.0046 | 0.653 | -0.0079 | 0.4656 |
| Glycine | rs2193619 | 2 | 211620445 | T | 0.838036 | 0.175935 | 7.95E-35 | 0.0065 | 0.7363 | 0.0215 | 0.2989 |
| Glycine | rs2287598 | 2 | 211523163 | G | 0.835212 | 0.102687 | 3.18E-11 | 0.0072 | 0.5234 | -0.0066 | 0.5821 |
| Glycine | rs2302908 | 2 | 211503736 | C | 0.883094 | 0.11793 | 8.42E-13 | -0.0051 | 0.752 | 0.0129 | 0.4446 |
| Glycine | rs2371011 | 2 | 211514492 | G | 0.361583 | 0.115609 | 3.33E-25 | 0.0039 | 0.7196 | -0.0016 | 0.8939 |
| Glycine | rs2371172 | 2 | 212105386 | C | 0.496487 | 0.066441 | 2.51E-09 | 0.0124 | 0.1218 | 0.0069 | 0.4164 |
| Glycine | rs2542929 | 2 | 211711593 | G | 0.719377 | 0.132802 | 2.94E-29 | -0.0133 | 0.1658 | 0.0046 | 0.6517 |
| Glycine | rs2719966 | 2 | 211840389 | A | 0.489672 | 0.148917 | 5.34E-46 | -0.0044 | 0.5927 | 0.004 | 0.6458 |
| Glycine | rs288096 | 2 | 210402536 | T | 0.861501 | 0.085455 | 2.81E-08 | 0.0024 | 0.8458 | -0.0014 | 0.9139 |
| Glycine | rs3748960 | 2 | 212247834 | C | 0.051241 | 0.149741 | 9.55E-09 | 0.0003 | 0.987 | -0.0133 | 0.4982 |
| Glycine | rs3770696 | 2 | 211310200 | T | 0.094218 | 0.104749 | 6.17E-09 | -0.0075 | 0.6037 | 0.0092 | 0.5527 |
| Glycine | rs4255905 | 2 | 211780849 | A | 0.844697 | 0.131777 | 1.47E-19 | -0.0093 | 0.451 | -0.0084 | 0.5233 |
| Glycine | rs4261668 | 2 | 211162312 | C | 0.223547 | 0.078386 | 1.43E-09 | 0.0062 | 0.5317 | 0.0015 | 0.8887 |
| Glycine | rs4299277 | 2 | 211726023 | G | 0.016033 | 0.538756 | 9.86E-23 | -0.0213 | 0.343 | 0.0131 | 0.6427 |
| Glycine | rs4461202 | 2 | 211795919 | T | 0.490968 | 0.168429 | 4.54E-52 | -0.0003 | 0.9709 | -0.0009 | 0.9176 |
| Glycine | rs4511662 | 2 | 211724050 | A | 0.565875 | 0.148916 | 2.09E-42 | 0.0141 | 0.1287 | 0.0035 | 0.7285 |
| Glycine | rs4520972 | 2 | 211765309 | C | 0.632746 | 0.108052 | 1.25E-22 | -0.0017 | 0.8403 | 0.0061 | 0.4939 |
| Glycine | rs4567871 | 2 | 211539302 | C | 0.852088 | 0.150523 | 4.56E-21 | -0.0069 | 0.5887 | -0.0153 | 0.2638 |
| Glycine | rs4600582 | 2 | 211785277 | C | 0.654452 | 0.099969 | 1.48E-19 | 0.0015 | 0.8623 | -0.0022 | 0.8117 |
| Glycine | rs4673545 | 2 | 211565535 | C | 0.376758 | 0.255046 | 2.09E-123 | -0.0206 | 0.02476 | 0.0098 | 0.326 |
| Glycine | rs4673553 | 2 | 211608379 | G | 0.517505 | 0.28572 | 6.64E-148 | -0.0155 | 0.06283 | 0.0126 | 0.1576 |
| Glycine | rs4673618 | 2 | 212310843 | G | 0.621245 | 0.073759 | 2.02E-10 | -0.0037 | 0.6649 | 0.0073 | 0.4239 |
| Glycine | rs6435477 | 2 | 209693319 | A | 0.26005 | 0.070359 | 4.17E-09 | -0.0056 | 0.5508 | 0.0015 | 0.8805 |
| Glycine | rs6714271 | 2 | 211764769 | A | 0.487655 | 0.148178 | 8.95E-45 | -0.0103 | 0.2105 | 0.0028 | 0.7506 |
| Glycine | rs6726552 | 2 | 211740016 | A | 0.584524 | 0.190055 | 1.12E-71 | 0.0006 | 0.9447 | 0.0047 | 0.6148 |
| Glycine | rs6743162 | 2 | 211765062 | T | 0.743257 | 0.146005 | 2.22E-34 | 0.0001 | 0.9919 | -0.0082 | 0.4305 |
| Glycine | rs6752652 | 2 | 211549240 | A | 0.482894 | 0.198557 | 4.69E-79 | -0.012 | 0.1344 | -0.0059 | 0.4978 |
| Glycine | rs6758081 | 2 | 211655965 | A | 0.80881 | 0.186546 | 7.95E-44 | -0.0076 | 0.4284 | -0.0061 | 0.5452 |
| Glycine | rs7574001 | 2 | 211569685 | T | 0.910024 | 0.176057 | 2.25E-20 | 0.0102 | 0.4354 | 0.0119 | 0.4063 |
| Glycine | rs7574280 | 2 | 211991553 | T | 0.34186 | 0.063193 | 1.07E-08 | 0.0112 | 0.1898 | -0.0052 | 0.5644 |
| Glycine | rs7577965 | 2 | 211904482 | T | 0.364956 | 0.101981 | 5.85E-18 | -0.0101 | 0.2655 | -0.0057 | 0.5594 |
| Glycine | rs7580498 | 2 | 211703869 | A | 0.691424 | 0.085986 | 2.26E-12 | 0.0018 | 0.8351 | 0.0086 | 0.3517 |
| Glycine | rs7585000 | 2 | 212264818 | C | 0.30632 | 0.068328 | 2.06E-09 | 0.0074 | 0.4091 | 0.0135 | 0.153 |
| Glycine | rs7593111 | 2 | 211724958 | C | 0.869109 | 0.129778 | 4.72E-17 | -0.0084 | 0.5401 | 0.0135 | 0.3567 |
| Glycine | rs7607199 | 2 | 211704032 | G | 0.695751 | 0.140809 | 2.26E-34 | -0.0005 | 0.9544 | 0.0095 | 0.3036 |
| Glycine | rs7608489 | 2 | 211699011 | A | 0.53495 | 0.134135 | 5.16E-33 | 0.007 | 0.3948 | -0.0043 | 0.6213 |
| Glycine | rs7684 | 2 | 211543147 | T | 0.648499 | 0.252989 | 2.14E-114 | -0.0124 | 0.1865 | -0.0033 | 0.7511 |
| Glycine | rs796224 | 2 | 211873239 | T | 0.369745 | 0.103024 | 3.41E-20 | -0.0127 | 0.1906 | 0.001 | 0.9264 |
| Glycine | rs888426 | 2 | 211713340 | T | 0.799091 | 0.166578 | 5.27E-37 | 0.0065 | 0.5165 | -0.003 | 0.7775 |
| Glycine | rs9646848 | 2 | 211669460 | T | 0.697419 | 0.159758 | 3.56E-44 | -0.005 | 0.5814 | -0.0041 | 0.6746 |
| Glycine | rs10106829 | 8 | 9184628 | A | 0.242797 | 0.087626 | 1.87E-11 | -0.0189 | 0.06745 | -0.0004 | 0.9711 |
| Glycine | rs330096 | 8 | 9171595 | C | 0.265249 | 0.075288 | 6.93E-10 | -0.0146 | 0.1916 | -0.0048 | 0.6941 |
| Glycine | rs6601299 | 8 | 9184691 | T | 0.141724 | 0.114869 | 3.72E-14 | -0.0256 | 0.07429 | 0.0032 | 0.8353 |
| Glycine | rs983309 | 8 | 9177732 | T | 0.162924 | 0.109592 | 3.69E-13 | -0.0148 | 0.2767 | -0.0034 | 0.8151 |
| Glycine | rs10739110 | 9 | 6697128 | T | 0.39034 | 0.079162 | 6.65E-12 | -0.0033 | 0.6993 | -0.0035 | 0.6981 |
| Glycine | rs10975136 | 9 | 5488594 | T | 0.283073 | 0.063525 | 4.41E-08 | -0.005 | 0.5725 | -0.0031 | 0.7428 |
| Glycine | rs10975159 | 9 | 5524571 | A | 0.044629 | 0.23657 | 1.42E-17 | 0.0001 | 0.9976 | 0.0051 | 0.8958 |
| Glycine | rs13289592 | 9 | 5966898 | C | 0.11606 | 0.121556 | 1.25E-12 | -0.0008 | 0.9524 | -0.0153 | 0.2748 |
| Glycine | rs13298772 | 9 | 5934989 | C | 0.053088 | 0.273079 | 3.74E-33 | 0.016 | 0.4019 | -0.0206 | 0.3171 |
| Glycine | rs13300552 | 9 | 6095799 | C | 0.06915 | 0.221241 | 1.37E-26 | 0.01 | 0.5879 | -0.023 | 0.244 |
| Glycine | rs2282162 | 9 | 6534466 | G | 0.53348 | 0.072139 | 1.48E-11 | -0.0084 | 0.3072 | -0.007 | 0.4212 |
| Glycine | rs2770761 | 9 | 7198855 | G | 0.038344 | 0.148142 | 3.88E-08 | -0.0271 | 0.2863 | -0.0104 | 0.703 |
| Glycine | rs369756 | 9 | 6146441 | T | 0.237429 | 0.072043 | 6.98E-09 | -0.0144 | 0.1549 | 0.0124 | 0.2474 |
| Glycine | rs450108 | 9 | 6153485 | C | 0.415981 | 0.06368 | 2.66E-09 | -0.0013 | 0.8775 | -0.0221 | 0.01318 |
| Glycine | rs4742211 | 9 | 6535500 | C | 0.184856 | 0.095054 | 4.06E-12 | -0.0055 | 0.5946 | -0.0168 | 0.1244 |
| Glycine | rs7032572 | 9 | 6172380 | G | 0.154515 | 0.110741 | 2.44E-14 | -0.0184 | 0.1062 | -0.0157 | 0.1905 |
| Glycine | rs7852365 | 9 | 6068910 | T | 0.144434 | 0.098228 | 2.27E-11 | -0.0004 | 0.9722 | -0.0016 | 0.8948 |
| Creatinine | rs12500824 | 4 | 77416627 | G | 0.65738 | 0.052862 | 3.84E-08 | -0.0072 | 0.3993 | 0.0034 | 0.7063 |
| Creatinine | rs2279463 | 6 | 160668389 | G | 0.110865 | 0.083336 | 3.24E-08 | 0.0355 | 0.004336 | -0.0315 | 0.0167 |
| Creatinine | rs7757336 | 6 | 160689558 | G | 0.13916 | 0.080168 | 1.46E-08 | 0.0204 | 0.07596 | -0.0222 | 0.06895 |
| Creatinine | rs17536527 | 15 | 45719187 | C | 0.435113 | 0.063465 | 5.41E-11 | -0.017 | 0.07976 | -0.0027 | 0.7992 |
| Creatinine | rs8037583 | 15 | 45546378 | T | 0.363879 | 0.055773 | 5.55E-09 | -0.0143 | 0.1023 | -0.005 | 0.5924 |
| Creatinine | rs950027 | 15 | 45801035 | C | 0.469892 | 0.052824 | 1.59E-08 | -0.006 | 0.4982 | 0.002 | 0.8323 |
| Creatinine | rs9806699 | 15 | 45740392 | A | 0.293605 | 0.068536 | 1.38E-11 | 0.0105 | 0.2633 | 0.0086 | 0.3887 |
| Creatinine | rs1990292 | 17 | 59444758 | G | 0.20606 | 0.069725 | 8.12E-09 | 0.016 | 0.114 | -0.0275 | 0.01189 |
| Creatinine | rs2286526 | 17 | 59472057 | C | 0.271962 | 0.065082 | 2.82E-09 | 0.0129 | 0.1549 | -0.0167 | 0.1084 |
| PC | rs10493322 | 1 | 62905893 | C | 0.730228 | 0.076821 | 0.000000027 | -0.0147 | 0.08917 | 0.0262 | 0.004101 |
| PC | rs964184 | 11 | 116648917 | G | 0.14413 | 0.100909 | 7.01E-09 | -0.0029 | 0.8061 | 0.0034 | 0.786 |
| PC | rs1077835 | 15 | 58723426 | G | 0.250724 | 0.18377 | 5.91E-37 | 0.0016 | 0.8744 | 0.018 | 0.09314 |
| PC | rs11632618 | 15 | 58724706 | A | 0.062544 | 0.177737 | 2.03E-10 | -0.0186 | 0.3136 | -0.0325 | 0.1239 |
| PC | rs11858164 | 15 | 58742731 | T | 0.475578 | 0.078666 | 9.67E-10 | -0.0189 | 0.04646 | -0.0289 | 0.005954 |
| PC | rs12905732 | 15 | 58683542 | C | 0.207477 | 0.089263 | 5.88E-09 | -0.0006 | 0.9542 | 0.0036 | 0.7443 |
| PC | rs166358 | 15 | 58680805 | A | 0.163819 | 0.129805 | 5.97E-15 | -0.0129 | 0.2833 | -0.0273 | 0.03208 |
| PC | rs17240832 | 15 | 58687370 | G | 0.176098 | 0.114819 | 1.63E-12 | -0.0155 | 0.1496 | 0.022 | 0.05273 |
| PC | rs17821316 | 15 | 58703069 | A | 0.499295 | 0.098791 | 2.51E-13 | -0.0046 | 0.6354 | -0.0128 | 0.2462 |
| PC | rs261333 | 15 | 58727279 | T | 0.494183 | 0.075535 | 2.33E-09 | -0.0133 | 0.1565 | -0.0134 | 0.1793 |
| PC | rs473224 | 15 | 58737341 | T | 0.146421 | 0.143634 | 2.4E-16 | -0.0096 | 0.4205 | -0.0226 | 0.07357 |
| PC | rs4775039 | 15 | 58670897 | G | 0.504641 | 0.085442 | 8.34E-10 | -0.0106 | 0.31 | 0.0068 | 0.5673 |
| PC | rs493258 | 15 | 58687880 | T | 0.498296 | 0.112299 | 1.36E-19 | -0.0042 | 0.6097 | -0.0076 | 0.3826 |
| PC | rs7170361 | 15 | 58718998 | C | 0.293535 | 0.140457 | 3.22E-24 | -0.0016 | 0.8782 | 0.0145 | 0.2017 |
| SM | rs12086676 | 1 | 55738663 | C | 0.81539 | 0.092269 | 8.18E-09 | 0.03 | 0.007284 | -0.0146 | 0.2194 |
| SM | rs7528419 | 1 | 109817192 | A | 0.786488 | 0.087711 | 4.99E-09 | -0.012 | 0.231 | -0.0235 | 0.02683 |
| SM | rs13392272 | 2 | 21217490 | T | 0.399678 | 0.070709 | 2.08E-08 | -0.0127 | 0.1274 | -0.0237 | 0.007141 |
| SM | rs1077835 | 15 | 58723426 | G | 0.249877 | 0.118363 | 3.34E-16 | 0.0016 | 0.8744 | 0.018 | 0.09314 |
| SM | rs16940170 | 15 | 58684282 | A | 0.159457 | 0.092467 | 4.31E-08 | -0.0172 | 0.1525 | -0.0288 | 0.02375 |
| SM | rs572410 | 15 | 58741384 | C | 0.281085 | 0.08682 | 3.05E-09 | -0.0098 | 0.3939 | -0.0246 | 0.05146 |
| SM | rs7170361 | 15 | 58718998 | C | 0.293136 | 0.086179 | 4.96E-10 | -0.0016 | 0.8782 | 0.0145 | 0.2017 |
| SM | rs17824149 | 17 | 4673185 | A | 0.877782 | 0.111584 | 3.59E-08 | -0.0191 | 0.4409 | -0.022 | 0.4218 |
| SM | rs7254892 | 19 | 45389596 | G | 0.975189 | 0.287476 | 1.17E-10 | 0.0222 | 0.3744 | -0.0114 | 0.6712 |
| TAG | rs1168041 | 1 | 62960250 | C | 0.722006 | 0.095641 | 3.4E-17 | -0.0155 | 0.09116 | 0.0304 | 0.001647 |
| TAG | rs12092541 | 1 | 62917913 | G | 0.834077 | 0.088551 | 5.64E-11 | -0.0203 | 0.0642 | 0.0217 | 0.06308 |
| TAG | rs10188514 | 2 | 21134064 | C | 0.326317 | 0.06674 | 3.03E-10 | 0.0109 | 0.2766 | 0.0078 | 0.4756 |
| TAG | rs13392272 | 2 | 21217490 | T | 0.412218 | 0.068882 | 1.31E-11 | -0.0127 | 0.1274 | -0.0237 | 0.007141 |
| TAG | rs1367117 | 2 | 21263900 | A | 0.287039 | 0.063968 | 9.07E-09 | -0.0105 | 0.2895 | -0.0126 | 0.2445 |
| TAG | rs1178979 | 7 | 72856430 | T | 0.828446 | 0.093267 | 1.13E-12 | -0.0082 | 0.4322 | -0.0075 | 0.4969 |
| TAG | rs4128744 | 8 | 19919655 | C | 0.89361 | 0.12404 | 3.29E-13 | -0.0153 | 0.215 | 0.0055 | 0.671 |
| TAG | rs6999158 | 8 | 19928013 | T | 0.655396 | 0.06457 | 2.81E-09 | -0.0144 | 0.125 | 0.0132 | 0.1858 |
| TAG | rs1729410 | 11 | 116665661 | G | 0.499179 | 0.066482 | 9.83E-11 | 0 | 1 | -0.001 | 0.9234 |
| TAG | rs180327 | 11 | 116623659 | C | 0.39629 | 0.077525 | 1.44E-14 | 0.0057 | 0.5046 | -0.0139 | 0.1278 |
| TAG | rs180360 | 11 | 116598988 | A | 0.689815 | 0.059102 | 3.04E-08 | 0.0006 | 0.9479 | -0.0095 | 0.3306 |
| TAG | rs486394 | 11 | 116526322 | C | 0.306893 | 0.082045 | 1.36E-13 | 0.0047 | 0.6425 | -0.0036 | 0.7419 |
| TAG | rs4938315 | 11 | 116731205 | G | 0.169279 | 0.074186 | 2.11E-08 | -0.0073 | 0.5641 | 0.0064 | 0.6349 |
| TAG | rs4938338 | 11 | 116972894 | A | 0.133506 | 0.079961 | 3.59E-08 | 0.0098 | 0.4713 | 0.0066 | 0.6475 |
| TAG | rs6589563 | 11 | 116590787 | A | 0.116905 | 0.095199 | 7.5E-10 | 0.0054 | 0.7203 | -0.011 | 0.4925 |
| TAG | rs6589567 | 11 | 116670676 | A | 0.155723 | 0.120194 | 3.38E-18 | -0.0036 | 0.7881 | -0.0101 | 0.4809 |
| TAG | rs964184 | 11 | 116648917 | G | 0.138247 | 0.232689 | 6E-61 | -0.0029 | 0.8061 | 0.0034 | 0.786 |
| TAG | rs10119 | 19 | 45406673 | A | 0.280337 | 0.064711 | 1.45E-08 | -0.0099 | 0.3713 | 0.0057 | 0.6376 |
| TAG | rs157580 | 19 | 45395266 | A | 0.700085 | 0.069404 | 1.5E-10 | -0.0024 | 0.7839 | 0.0019 | 0.8388 |
| TAG | rs2304128 | 19 | 19746151 | G | 0.924838 | 0.123534 | 1.65E-10 | -0.0236 | 0.1305 | 0.0268 | 0.1011 |
| TAG | rs405697 | 19 | 45404691 | G | 0.775919 | 0.077002 | 3.65E-10 | -0.0106 | 0.352 | -0.0148 | 0.2373 |
| TAG | rs439401 | 19 | 45414451 | C | 0.691135 | 0.09236 | 1.4E-16 | -0.0157 | 0.1056 | 0.001 | 0.9226 |
| TAG | rs4608457 | 19 | 20217169 | C | 0.95533 | 0.155523 | 1.45E-08 | 0.014 | 0.8926 | N.A | N.A |

EA=effect allele. EAF=effect allele frequency. BMD=bone mineral density. FN=femoral neck. LS=lumbar spine. PC=phosphatidylcholine. SM=Sphingomyelin. TAG=Triacylglycerol. N.A.=not available

**Supplementary Table 8.** Power calculation for two-sample Mendelian randomization with fixed α=0.05

| **Exposure** | **Outcome** | **n.exposure** | **n.outcome** | **Variance of exposure** | **Variance of outcome** | **n.IV** | **r^2^** | | | **Causal effect** | **Estimated power** | | |
| --- | --- | --- | --- | --- | --- | --- | --- | --- | --- | --- | --- | --- | --- |
| Leucine | FN-BMD | 24728 | 32961 | 0.050 | 0.022 | 4 | 0.8% | 0.64% | 0.4% | -0.028 | 0.09 | 0.08 | 0.07 |
|  | LS-BMD | 24728 | 32961 | 0.050 | 0.045 | 4 | 0.8% | 0.64% | 0.4% | 0.071 | 0.18 | 0.16 | 0.12 |
| Glycine | FN-BMD | 18734 | 32961 | 0.118 | 0.022 | 85 | 12.49% | 10% | 8.5% | -0.033 | 0.99 | 0.97 | 0.94 |
|  | LS-BMD | 18734 | 32961 | 0.118 | 0.045 | 85 | 12.49% | 10% | 8.5% | -0.013 | 0.22 | 0.18 | 0.16 |
| Creatinine | FN-BMD | 24810 | 32961 | 0.024 | 0.022 | 9 | 0.93% | 0.74% | 0.9% | 0.074 | 0.22 | 0.18 | 0.21 |
|  | LS-BMD | 24810 | 32961 | 0.024 | 0.045 | 9 | 0.93% | 0.74% | 0.9% | 0.146 | 0.36 | 0.30 | 0.35 |
| PC | FN-BMD | 13542 | 32961 | 0.058 | 0.022 | 14 | 4.26% | 3.41% | 1.4% | -0.066 | 0.93 | 0.87 | 0.51 |
|  | LS-BMD | 13542 | 32961 | 0.058 | 0.045 | 14 | 4.26% | 3.41% | 1.4% | -0.136 | 0.99 | 0.99 | 0.81 |
| SM | FN-BMD | 13476 | 32961 | 0.071 | 0.022 | 9 | 3.92% | 3.14% | 0.9% | -0.017 | 0.16 | 0.14 | 0.07 |
|  | LS-BMD | 13476 | 32961 | 0.071 | 0.045 | 9 | 3.92% | 3.14% | 0.9% | -0.145 | 0.99 | 0.99 | 0.77 |
| TAG | FN-BMD | 21545 | 32961 | 1.762 | 0.022 | 23 | 4.33% | 3.46% | 2.3% | -0.065 | 1 | 1 | 1 |
|  | LS-BMD | 21545 | 32961 | 1.762 | 0.045 | 22 | 4.33% | 3.46% | 2.2% | -0.070 | 1 | 1 | 1 |

n.exposure=sample size for exposure. n.outcome=sample size for outcome. n.IV=number of instrumental variables. r^2^=proportion of variance of exposure attributed to IVs; the first column is based from the original GWAS paper; the second column is 80% of the first columns since we selected the variants with $p<5\times{10}^{-8}$; the third column is assumed that each genome-wide significant variant explains 0.01% of the variance. The three columns under Estimated power are corresponding to the three settings for r_­_^2^. BMD=bone mineral density. FN=femoral neck. LS=lumbar spine. PC=phosphatidylcholine. SM=Sphingomyelin. TAG=Triacylglycerol.

Note: Power calculation were conducted using Deng et al.’s code ^(1)^. Causal effect refers to the change of outcome for each standard deviation change in exposure. Sample sizes of exposure and outcome are given by the GWAS papers ^(2,3)^. Variance of exposure and outcome are estimated from our study data. Proportion of exposure variance explained by the IVs are given by the GWAS paper ^(2)^. Since we only selected a subset of variants which are genome-wide significant, we also included two other settings for r^2^: the first one is 80% of given proportion; the second one is assumed that each genome-wide significant variant explains 0.01% of the variance.
